# COVID-19 DISEASE IN PEDIATRIC EMERGENCY ROOM: THE DILEMMA OF CYCLE THRESHOLD VALUE

**DOI:** 10.1101/2022.06.23.22276825

**Authors:** Demet Timur, Uğur Demirpek, Başak Ceylan Demirbaş, Esra Türe, Muhammed Furkan Korkmaz, Ahmet Timur

## Abstract

**BACKGROUND:** The havoc of this SARS-CoV-2 pandemic was being distributed unequally. Children, of all ages, and in all countries, are being affected. Testing for infection with SARS-CoV-2, helps to determine what interventions may need to be put in place to control the spread of disease within a community. A PCR test for COVID-19 is a test used to diagnose children infected with SARS-CoV-2, the virus that causes COVID-19. Cycle threshold (Ct) is a semi-quantitative value that tells us approximately how much viral genetic material is in the sample following testing by RT-PCR. Our aim was to evaluate how RT-PCR Ct values among children with confirmed SARS-CoV-2 compared with clinical, laboratory and demographic data.

**MATERIALS AND METHODS:** In the study, demographic, laboratory, radiological and clinical characteristics and the effect of Ct value of patients with positive Covid-19 PCR test who applied to the Emergency Pediatric Service with the suspicion of infectious disease between May 3,2020 and August 3, 2020 were retrospectively examined.

**RESULTS:** There was no statistical significance between the patient’s hospitalization status, admission symptoms other than fever, and laboratory parameters and the mean Ct value. It was determined that the mean Ct value of the patients who had symptoms at the time of admission to the hospital was statistically significantly lower.

**CONCLUSION:** In this study, symptomatic patients had lower Ct than asymptomatic patients; this reflects the clinical impact of increased viral load. In our study, the low Ct values in symptomatic patients and higher values in asymptomatic patients; reflects the importance of the effect on the clinic with the increase of viral load. In evidence-based medicine practices, it will be useful to check the compatibility of complaints and findings with laboratory data while evaluating patients. In addition, if the patient is symptomatic and has a high ct value, co-infections should be considered.

## INTRODUCTION

Many countries around the world have reduced or abolished prevention measures against COVID-19 infection. However, the COVID-19 pandemic has not ended, and it tends to continue its existence at different intensities.The hunch and analysis of all processes and dynamics to reduce the risk of recurrence of the disease will guide measures and work to be carried out in the future. Given the continued pandemic of Covid-19 and the rapid growth of research on this topic; the study of pediatric patients is challenging and indispensable. The clinical course of Covid-19 infections is milder in children than in adult patients[1]. However, in the medium to long term, it can also cause considerable complications in children as well. Furthermore, children may not have symptoms even during the acute disease. People with COVID-19 infections can have a wide range of symptoms, which requires differential diagnosis. Symptoms such as headache, sore throat, running nose, fever, cough are frequently observed. Though, few children may have respiratory problems and require intensive care; the COVID-19 infection causes children’s deaths worldwide. However, child mortality are often associated with conditions that are related to non-COVID-19 diseases. Besides, COVID-19 disease affects systems other than the respiratory system. Thus, children may develop a multisystem inflammatory syndrome called MIS-C within 3 to 6 weeks of COVID-19 diagnosis[2, 3].

The Real-time PCR test is the gold standard test for the diagnosis of COVID-19 infection. During test reaction fluorescence signals are generated depending on target gene amplification. The number of cycles required to detect these fluorescent signals is called the **Cycle threshold** ‘Ct’. Cycle threshold (Ct) is a semi-quantitative value that determines approximately how much viral genetic material is in the sample. The cycle threshold value and viral load are inversely proportional to each other. [4]. A low Ct indicates a high concentration of viral genetic material; vice versa, a high Ct indicates a low concentration of viral genetic material. The Ct value is affected by many factors.Those factors could be classified into two basic categories; respectively, endogenous and exogenous factors. In this study, we aim to analyze laboratory data and clinical characteristics of children with Covid-19 to determine whether Ct values affect diagnostic and follow-up parameters.

## MATERIALS AND METHODS

The study was conducted as a retrospective observational cohort study in the Emergency Department and the PediatricWards. From May 3, 2020 to August 3, 2020; 244 patients under the age of 18 with positive results of COVID-19 real-time reverse transcription polymerase chain reaction (RT-PCR) were included in the study. After the approvals of the local ethics council and the Turkish Ministry of Health, patient files and hospital automation system data were retrospectively reviewed and recorded. Clinical and epidemiological data on age, gender, symptoms at the time of presentation (fever, cough, myalgia, sore throat, headache, nausea, vomiting, diarrhea, loss of smell-taste, abdominal pain, dyspnea), laboratory results and medical history have been recorded.

Patients with negative COVID-19 RT-PCR results, aged over 18 years, or whose data were lacking were not included in the study. Combined oropharyngeal and nasopharyngeal samples from patients were sent to the laboratory in a virus transport medium at 4°C. The samples were analyzed using Bio-Speedy SARS-CoV-2 kit (Bioeksen, Turkiye) in a real-time Rotor-Gene Q PCR device (Qiagen, Germany) after extraction of nucleic acid. The Ct values of patients who had positive PCR results were recorded and statistically evaluated.

Clinical laboratory tests and C-reactive protein (CRP) were measured in a Cobas C 702 (Roche Diagnostics, Germany) analyzer from the sera of the patients. Prothrombin time (PT), activated partial thromboplastin time (aPTT) and D-dimer levels were measured with Cobas T 711 (Roche Diagnostics, Germany) analyzer. Complete blood count was analyzed on Sysmex XN-9100 hematology analyzer (Sysmex Corporation, Japan).

### Statistical analysis

Data was analyzed by using SPSS Statistics for Windows, Version 20.0. The expression n (%)has been used for categorical variables, and the mean ± SD (standard deviation) in accordance with the normal distribution continuous variables; in the case of compliance was not achieved, the median values (lower / higher limit) were used. Descriptive analyzes were used to analyze the distribution and frequency of data, and Chi-Square tests were used to compare two independent groups in categorical data. Shapiro-Wilk test was used to test analysis of normality. The student-t test was used to compare two independent groups with the normal distribution. The Mann-Whitney U test was used to compare two independent groups. The significance level was accepted as <0.05 in all statistical analyses.

## RESULTS

The patients who were admitted to the pediatric emergency department of our hospital and deemed positive for the COVID-19 RT-PCR test from May 3, 2020 to August 3, 2020 were 244 patients. Of these patients, 129 (52.9%) were male and 115 (47.1%) were female. The male/female ratio was 1.12 (129/115). The mean age of all patients was 8.79±5.52, while it was 9.19±5.83 for girls and 8.43±5.23 for boys. The demographic characteristics of the patients and their distribution by age groups are shown in Table 1.

**TABLE 1.** Demographic characteristics of patients and distribution by age groups.

|  | **n** | **Ratio (%)** |
| --- | --- | --- |
| **Sex** |  |  |
| Boys | 129 | 52,9 |
| Girls | 115 | 47,1 |
| **Age** |  |  |
| 0-4 | 71 | 29,1 |
| 5-9 | 59 | 24,2 |
| 10-14 | 63 | 25,8 |
| 15 and over | 51 | 20,9 |
| **Patient Group** |  |  |
| Inpatient / hospitalized | 48 | 19,7 |
| Outpatient | 196 | 80,3 |
| **Clinical Characteristics** |  |  |
| Symptomatic | 31 | 12,7 |
| Asymptomatic | 213 | 87,3 |

As the COVID-19 contact stories were questioned, it was seen that 239 (98%) had a contact history, only 5 (2%) did not declare a contact history. When the patients’ admission to the hospital were asked; It was observed that 213 (87.3%) had at least one symptom, 31 (12.7%) had no complaints and applied to the hospital due to contact history. When the symptoms of the patients were examined at the admission, it was found that the most common complaint was fever (n:141, 66.2%), followed by cough and myalgia.The distribution of patients according to symptoms at the time of presentation and the duration of symptoms are shown in Table 2.

**TABLE 2.**
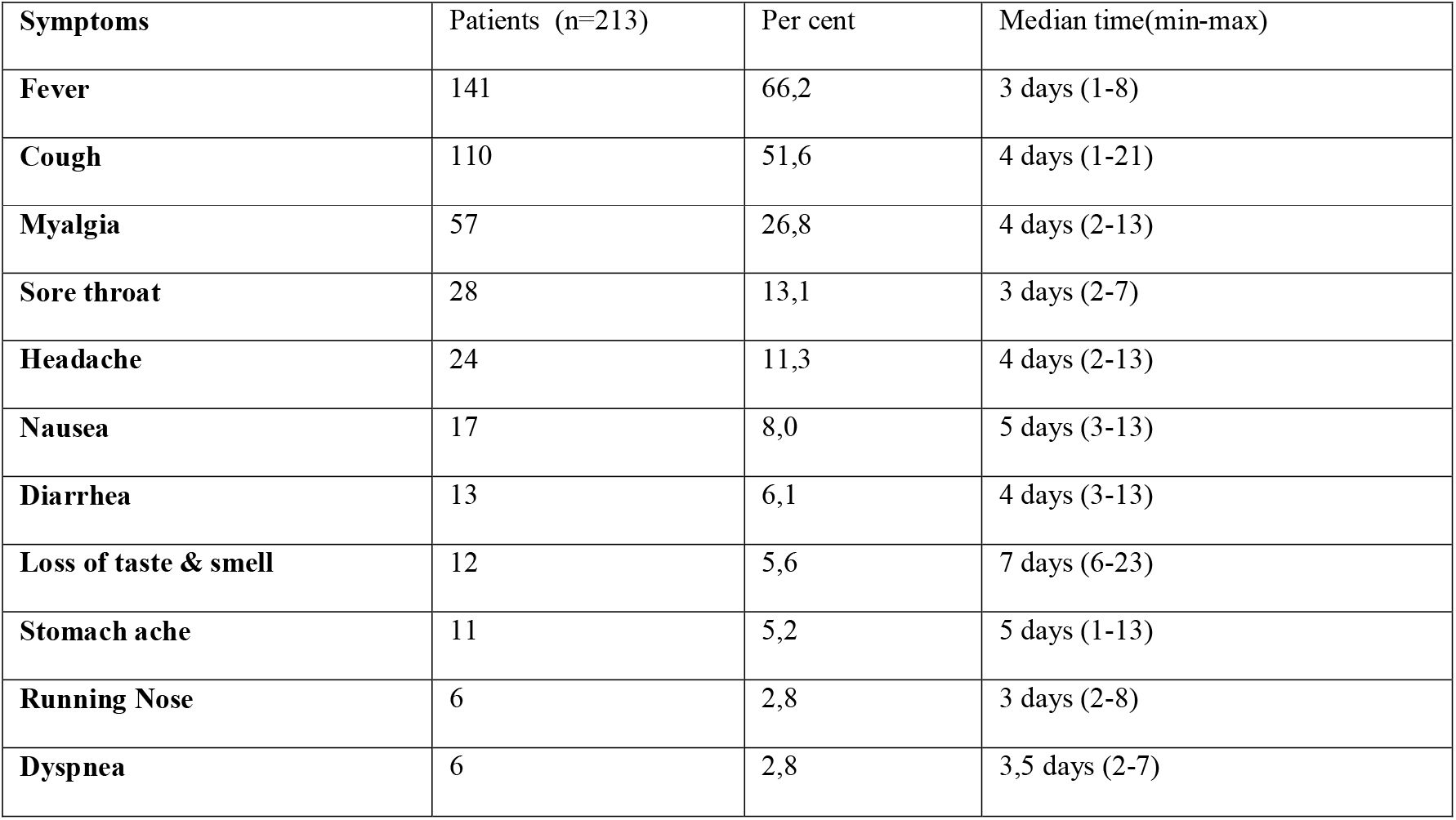
Distribution of symptoms at presentation and duration.

It was determined that 48 (19.7%) of 244 patients were hospitalized. When hospitalized patients were compared in terms of age groups, 22 (45.8%) of 48 patients were found to be between the ages of 0-4 (p=0.012). When inpatients and outpatients were compared according to the presenting symptoms, it was found that the patients with diarrhea (53.8%) were hospitalized more (p=0.001). No statistical significance was found as inpatients and outpatients were compared according to gender, contact history, and other clinical features(p>0.05). Table 3 shows the distribution of inpatients and outpatients by sex,age groups and symptoms.

**TABLE 3.**
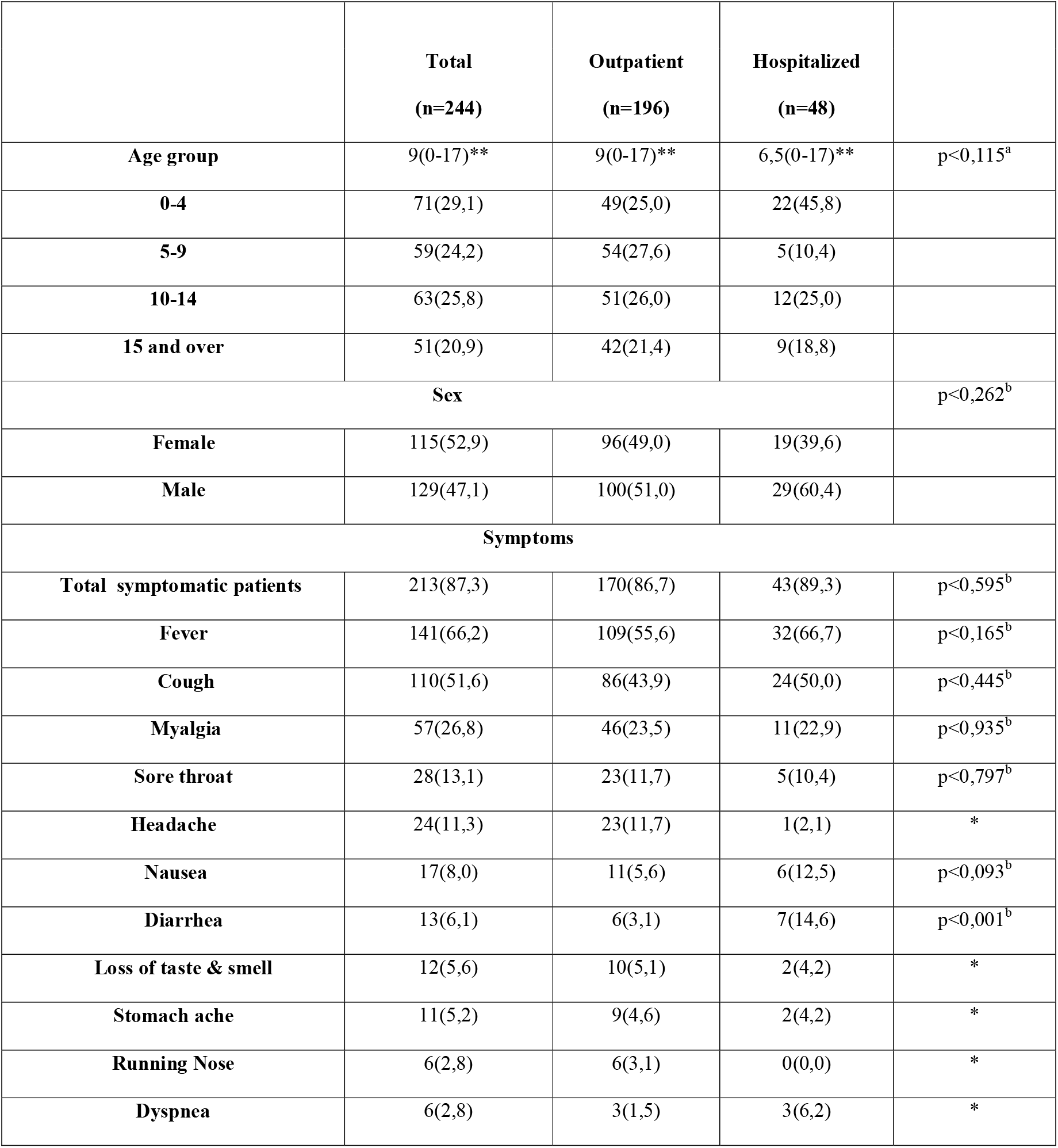
The distribution of inpatients and outpatients by sex, age groups and symptoms.

As the radiological findings of the 244 pediatric patients were examined; chest X-ray results were normal in 207 patients, pathological findings were detected in 37 patients. Thorax CT was performed in 18 patients and pathological findings were detected in 3 patients.

In this study, it was found that the mean COVID-19 RT-PCR Ct value of all patients was 22.35±4.47. It was determined that the mean Ct value of those with symptoms at the time of admission was 21.94±5.37, and 25.15±5.45 in those without symptoms. It was determined that the mean Ct value of the patients who had symptoms at the time of admission to the hospital was statistically significantly lower (p=0.002). When the mean Ct values were compared according to the symptoms at presentation, it was found that the mean Ct values of those who had fever at admission were statistically significantly lower than those who did not have fever (20.65±4.87, 24.67±5.28, respectively). While the mean Ct value of the boys was 22.22±5.1, it was 22.48±5.88 for the girls. It was determined that the mean Ct value did not differ statistically significantly according to gender (p>0.05). The mean Ct value of inpatients was 22.33±5.55, and it was 22.40±5.19 in outpatients. There was no statistical significance between the patient’s hospitalization status, admission symptoms other than fever, and the mean Ct value (p>0.05). The relationship between the laboratory parameters of the patients and the hospitalization status is shown in Table 4, and the relationship with the Ct value is shown in Table 5.

**TABLE 4.** The relationship between the laboratory parameters and the inpatients.

|  | **Total**<br>**(n=244)** | **Outpatient**<br>**(n=196)** | **Hospitalized**<br>**(n=48)** |  |
| --- | --- | --- | --- | --- |
| **WBC <4** | 20(8,2) | 17(8,7) | 3(6,2) | $p<0,583$ |
| **Lymphocyte<0,8** | 7(2,9) | 4(2,0) | 3(6,2) | $p<0,117$ |
| **AST>40** | 22(9,0) | 12(6,1) | 10(20,8) | $p<0,001$ |
| **ALT>40** | 0 | 0 | 0 |  |
| **LDH>300** | 27(11,1) | 16(8,2) | 11(22,9) | $p<0,003$ |
| **PT>16** | 0 | 0 | 0 |  |
| **D-dimer** | 21(8,6) | 9(4,6) | 12(25,0) | $p<0,001$ |
| **ESR>20** | 48(19,7) | 36(18,4) | 12(25,0) | $p<0,300$ |
| **CRP>8** | 21(8,6) | 14(7,1) | 7(14,6) | $p<0,099$ |
| **Procalcitonin>0,5** | 2(0,8) | 1(0,5) | 1(2,1) | $p<0,279$ |

WBC:White Blood Cell, AST: Aspartate Transaminase, ALT: Alanine Transaminase, LDH: Lactate Dehydrogenase, PT: Prothrombin time, ESR: Erythrocyte sedimentation rate, CRP: C-Reactive protein.

**TABLE 5.** The relationship between the laboratory parameters and the Ct values.

|  | Toplam(n=244) | <22,35(n=133) | >22,35(n=111) | p |
| --- | --- | --- | --- | --- |
| **WBC <4** | 20(8,2) | 10(7,5) | 10(9,0) | 0,673 |
| **Lymphocyte<0,8** | 7(2,9) | 4(3,0) | 3(2,7) | 0,887 |
| **AST>40** | 22(9,0) | 13(9,8) | 9(8,1) | 0,651 |
| **ALT>40** | 0 | 0 | 0 |  |
| **LDH>300** | 27(11,1) | 17(12,8) | 10(9,0) | 0,350 |
| **PT>16** | 0 | 0 | 0 |  |
| **D-dimer** | 21(8,6) | 12(9,0) | 9(8,1) | 0,800 |
| **ESR>20** | 48(19,7) | 24(18,0) | 24(21,6) | 0,484 |
| **CRP>8** | 21(8,6) | 13(9,8) | 8(7,2) | 0,476 |
| **Procalcitonin>0,5** | 2(0,8) | 2(1,5) | 0 | 0,195 |

WBC:White Blood Cell, AST: Aspartate Transaminase, ALT: Alanine Transaminase, LDH: Lactate Dehydrogenase, PT: Prothrombin time, ESR: Erythrocyte sedimentation rate, CRP: C-Reactive protein.

## DISCUSSION

The SARS-COV-2 pandemic has affected the whole world and is still active. Although vaccination studies and infection control measures have been successful, it is not clear where the epidemic will be headed in the near future. In this study, we aimed to contribute to the literature on pediatric SARS-COV-2 infection by the analysis of the relationship between the Ct value and the patient data. All patients involved in this study, including those with comorbidities, recovered well and did not need intensive care support during their treatment. Most of the studies hint that transmission of Severe Acute Respiratory Syndrome Coronavirus 2 (SARS-CoV-2) occurs within household contacts[6-8]. In our study, household contacts were found to be 98% in accordance with the literature. The rate of male patients was noted 53% in our study, it was found to be between 45-55% in similar studies [7-12].

In our study, it was found that the proportion of asymptomatic patients was 13%, and this rate varied from 6 to 48% in different studies. [7-13]. When analyzing the symptoms, the most common symptoms were fever and cough, similar to our study. [7-16]. In a study assessing data for 1734 children aged 5-17 in the United Kingdom, however, it was found that the most common symptoms were headache (62%) and fatigue(55%) [17]. SARS-CoV-2 infections in children have different paths and are less severe than in adults in terms of clinical manifestations [18]. In a Chinese study, diarrhea was observed only in severe patients [19], while some studies pointed out that it was more common in children with mild COVID-19 [8, 9]. As compared to patients based on presenting symptoms, 53.8% of patients with diarrhea were hospitalized in our study.

Cui et al. in their study, found the rate of patients with normal WBC values was 69%. [12]. Similarly, in our study, this rate was found to be 84%. Lympopenia, another laboratory finding, was 14-36% in a few studies [8, 9, 12, 20]. Yayla et al. in their study, lymphopenia was observed at a rate of 14% [9]. In our study, the rate of lymphopenia was quite low, which was statistically insignificant.

Since previous studies have shown that leukocyte indexes have a wide range of distribution in children compared to adults with COVID-19; monitoring Levels Of CRP, LDH, and PCT appears to be more important for showing how the patients progress, [21]. In a study by Yildiz et al., CRP and PCT levels in symptomatic children were also significantly higher[22]. Some studies have shown that the elevation of the CRP is 18-19% and the elevation of the LDH is 28-29% [8, 12]. Yayla et al. In their study, CRP and LDH values were normal in most of the patients. However, an increase in LDH was seen in severe patients in the study [9]. Elevated PCT levels was found to be 36% of the patients in a study conducted by Cui et al. Whereas, Yayla et al Procalcitonin levels were normal in most patients [8, 9, 12]. In our study, CRP, PCT and LDH were found to be normal in most of the patients. We think that the lack of clinically serious patients in this study was the main cause and one of the limitations for obtaining these results. Yildiz et al. found the N/L ratio to be significantly higher in symptomatic children in their studies [22]. High neutrophil to lymphocyte ratio (NLR) is envisaged as an objectionable prognostic factor for COVID-19 infection in many researches [22, 23]. In our study, the N/L ratio was found to be higher in symptomatic patients. It is recommended to investigate the possibility of COVID-19 in patients with symptoms of influenza infection who do not have high AST values [24]. In our study, the rate of patients with elevated AST was very low, 9%. However, it is noteworthy that it is significantly higher in hospitalized patients.

Abnormal findings were found at a rate of 33-54% in studies examining the results of chest radiography [8, 16]. In our study, this rate was found to be 14%. In a pediatric study conducted in our country, Boncuoglu et al. abnormal CT findings and elevated CRP have been shown to accompany patients [25]. In our study, the number of patients with abnormal CT results were statistically insignificant.

Chung et al., found out that symptomatic and asymptomatic patients were evaluated independently, the mean Ct value of children in both groups was found to be similar to adults [26]. When compared to adults, symptomatic children with similar Ct values have higher viral loads even in the initial period of the disease. It can explain the relevant role of children in the transmission of SARS-CoV-2. [27]. Fox-Lewis et al. proposed in their study that Ct values should not be used as absolute markers of the period after infection or to exclude infection when the date of symptom onset is not available. [28]. As symptomatic and asymptomatic pediatric cases researched thoroughly, no significant differences were found between Ct values[6, 11]. Despite; it is suggested that children are not the main drivers of the transmission of SARS-CoV-2 because compared to adults, children who had tested positive for SARS-CoV-2 were less likely to grow virus in culture and had higher cycle thresholds and lower viral concentrations [29]. Inconsistently, the fact that symptomatic pediatric patients have lower Ct values than asymptomatic patients indicates that the viral load is higher in these patients [13, 26, 30]. In our study, the mean Ct value of patients with symptoms at admission was found to be statistically significantly lower. In some studies, the children’s age differences were noted. Children under five years of age have a statistically significantly lower Ct value, indicating a higher viral load in patients in this age group [13, 31]. It is believed that the patients’ Ct value will guide the decision-making of the clinical processes.[32]. It is noted that viral load is important in determining the course of the disease in studies with adults.In addition, it was also observed that viral burden persists long-term high levels in severely ill patients [19, 30]. The Ct value, a semi-quantitative indicator of viral genetic material concentrations, is considerably relative in COVID-19 RT-PCR diagnostic studies and must be interpreted based on various factors. From a laboratory perspective, Ct values should only be reported and applied for clinical interpretation and action where the linearity, limit of detection and standard quantification curves are assured. The interpretation of a single positive Ct value for the treatment of infectious disease, prognosis, infection,or as a recovery indicator must be done in the context of clinical history. Low Ct values (high viral load) are more likely to show acute disease and infectiousness. High Ct values (low viral load) may be due to a variety of clinical scenarios to reduce the risk of infection, but interpretation requires clinical context. Serial Ct values are more profitable for interpretation, but are frequently carried out only in hospital settings for clinical management purposes rather than for infection control issues [33].

## CONCLUSION

Ct value results should be used with clinical context for decision making about a patient’s infectivity. Many laboratories use different test methods and some may use more than one, hence, Ct values should not be directly compared between different assay types. PCR tests have made a significant contribution to the diagnostic process of COVID-19 and to distinguish between other challenging viral infections in children. In our study, Ct values in symptoms are lower than in non-symptomatic patients, which shows the clinical effects of increasing viral load. If the patient is symptomatic and has a high ct value; newer variants or co-infections, especially viral agents such as influenza viruses, should be considered. There is an indispensable need for further research to understand the impact of Ct values on laboratory results and clinical impact in children.

## Data Availability

All data produced in the present work are contained in the manuscript

https://bursasehir.saglik.gov.tr/?_Dil=2

## REFERENCES

1. Badal, S., et al., Prevalence, clinical characteristics, and outcomes of pediatric COVID-19: A systematic review and meta-analysis. Journal of Clinical Virology, 2021. 135: p. 104715.

2. Henderson, L.A. and R.S. Yeung, MIS-C: early lessons from immune profiling. Nature Reviews Rheumatology, 2021. 17(2): p. 75–76.

3. Sancho-Shimizu, V., et al., SARS-CoV-2–related MIS-C: A key to the viral and genetic causes of Kawasaki disease? Journal of Experimental Medicine, 2021. 218(6): p. e20210446.

4. Gontla, V., et al., “prognostic value of cycle threshold” in COVID-19 confirmed patients. Indian Journal of Critical Care Medicine, 2021: p. S55–S55.

5. Korkmaz, M.F., et al., The epidemiological and clinical characteristics of 81 children with COVID-19 in a pandemic hospital in Turkey: an observational cohort study. Journal of Korean medical science, 020. 35(25).

6. Chu, M.A., et al., Viral load and rebound in children with coronavirus disease 2019 during the first outbreak in Daegu city. Clinical and experimental pediatrics, 2021. 64(12): p. 652.

7. Sim, J.Y., et al., Characteristics, contacts, and relative risk of SARS-CoV-2 infection among children during school closures. Journal of Microbiology, Immunology and Infection, 2021.

8. Yayla, B.C.C., et al., Characteristics and management of children with COVID-19 in Turkey. Balkan Medical Journal, 2020. 37(6): p. 341.

9. Yayla, B.C.C., et al., Characteristics and management of children with COVID-19 in a tertiary care hospital in Turkey. Clinical pediatrics, 2021. 60(3): p. 170–177.

10. Sahin, A., et al., Pediatric Patients with COVID-19: A Retrospective Single-Center Experience. The Medical Bulletin of Sisli Etfal Hospital, 2022. 56(1): p. 62–69.

11. Peaper, D.R., et al., Severe acute respiratory syndrome coronavirus 2 testing in children in a large regional US health system during the coronavirus disease 2019 pandemic. The Pediatric Infectious Disease Journal, 2021. 40(3): p. 175–181.

12. Cui, X., et al., A systematic review and meta□analysis of children with coronavirus disease 2019 (COVID□19). Journal of medical virology, 2021. 93(2): p. 1057–1069.

13. Strutner, J., et al., Comparison of Reverse-Transcription Polymerase Chain Reaction Cycle Threshold Values From Respiratory Specimens in Symptomatic and Asymptomatic Children With Severe Acute Respiratory Syndrome Coronavirus 2 Infection. Clinical Infectious Diseases, 2021. 73(10): p. 1790–1794.

14. Berksoy, E., et al., Clinical and laboratory characteristics of children with SARS□CoV□2 infection. Pediatric Pulmonology, 2021. 56(12): p. 3674–3681.

15. Alp, E.E., et al., Evaluation of Patients with Suspicion of COVID-19 in Pediatric Emergency Department. The Medical Bulletin of Sisli Etfal Hospital, 2021. 55(2): p. 179.

16. Sedighi, I., et al., A multicenter retrospective study of clinical features, laboratory characteristics, and outcomes of 166 hospitalized children with coronavirus disease 2019 (COVID□19): A preliminary report from Iranian Network for Research in Viral Diseases (INRVD). Pediatric pulmonology, 2022. 57(2): p. 498–507.

17. Molteni, E., et al., Illness duration and symptom profile in symptomatic UK school-aged children tested for SARS-CoV-2. The Lancet Child & Adolescent Health, 2021. 5(10): p. 708–718.

18. Garazzino, S., et al., Multicentre Italian study of SARS-CoV-2 infection in children and adolescents, preliminary data as at 10 April 2020. Eurosurveillance, 2020. 25(18): p. 2000600.

19. Zheng, S., et al., Viral load dynamics and disease severity in patients infected with SARS-CoV-2 in Zhejiang province, China, January-March 2020: retrospective cohort study. bmj, 2020. 369.

20. Alkan, G., et al., Evaluation of hematological parameters and inflammatory markers in children with COVID-19. Irish Journal of Medical Science (1971-), 2021: p. 1–9.

21. Henry, B.M., et al., Laboratory abnormalities in children with mild and severe coronavirus disease 2019 (COVID-19): a pooled analysis and review. Clinical biochemistry, 2020. 81: p. 1–8.

22. Yildiz, E., et al., High Neutrophil/Lymphocyte Ratios in Symptomatic Pediatric COVID-19 Patients. Journal of the College of Physicians and Surgeons--pakistan: JCPSP, 2021. 31(7): p. 93–98.

23. Attia Bari, A.C. and N.S. Iqbal Bano, Is leukopenia and lymphopenia a characteristic feature of COVID-19 in children? Pakistan Journal of Medical Sciences, 2021. 37(3): p. 869.

24. Liu, X.P., et al., Comparison of laboratory data between children with COVID□19 and influenza. The Kaohsiung Journal of Medical Sciences, 2021. 37(2): p. 158.

25. Böncüoğlu, E., et al., Can laboratory findings predict pulmonary involvement in children with COVID□19 infection? Pediatric Pulmonology, 2021. 56(8): p. 2489–2494.

26. Chung, E., et al., Comparison of symptoms and RNA levels in children and adults with SARS-CoV-2 infection in the community setting. JAMA pediatrics, 2021. 175(10): p. e212025–e212025.

27. Polese-Bonatto, M., et al., Children Have Similar Reverse Transcription Polymerase Chain Reaction Cycle Threshold for Severe Acute Respiratory Syndrome Coronavirus 2 in Comparison With Adults. The Pediatric infectious disease journal, 2021. 40(11): p. e413.

28. Fox-Lewis, A., et al., SARS-CoV-2 viral load dynamics and real-time RT-PCR cycle threshold interpretation in symptomatic non-hospitalised individuals in New Zealand: a multicentre cross sectional observational study. Pathology, 2021. 53(4): p. 530–535.

29. Bullard, J., et al., Infectivity of severe acute respiratory syndrome coronavirus 2 in children compared with adults. Cmaj, 2021. 193(17): p. E601–E606.

30. Liu, Y., et al., Viral dynamics in mild and severe cases of COVID-19. The Lancet infectious diseases, 2020. 20(6): p. 656–657.

31. Heald-Sargent, T., et al., Age-related differences in nasopharyngeal severe acute respiratory syndrome coronavirus 2 (SARS-CoV-2) levels in patients with mild to moderate coronavirus disease 2019 (COVID-19). JAMA pediatrics, 2020. 174(9): p. 902–903.

32. Tom, M.R. and M.J. Mina, To interpret the SARS-CoV-2 test, consider the cycle threshold value. Clinical Infectious Diseases, 2020.

33. Katsuta T, Shimizu N, Okada K, et al., The clinical characteristics of pediatric coronavirus disease 2019 in 2020 in Japan. Pediatr Int. 2022 Jan;64(1):e14912. doi: 10.1111/ped.14912.

